# Algorithmic Virtual Reality Reduces Parkinsonian Tremor

**DOI:** 10.1101/2023.02.05.22283323

**Authors:** John Cornacchioli, Alec Galambos, Stamatina Rentouli, Robert Canciello, Roberta Marongiu, Daniel Cabrera, eMalick G. Njie

## Abstract

Parkinson’s disease (PD) is one of the most prevalent neurodegenerative disorders, affecting approximately 6-7 million patients worldwide. Involuntary hand tremor (IHT), one of the cardinal motor symptoms of PD, is extremely debilitating to patients reducing their quality of life. In this study, we combined neuroscience together with the nascent field of medical virtual reality (VR). Our goals were to 1) generate technology to enable PD patients to leapfrog the real life difficulties of living with IHT by entering VR metaverse worlds in which they are tremor-free and can function at new heights, 2) to determine whether lack of tremors in VR modifies central biofeedback mechanisms to non-invasively reduce tremors in real life. We thus generated algorithms that adjusted the moving average centroid of digital hands to stabilize tremors in VR. We implemented these algorithms in the Oculus Rift VR system and enrolled nine subjects with PD in a trial that yielded several important observations. First, we established the Oculus Rift as a potent measurement device for IHT. Secondly, we determined parkinsonian tremors can be abolished in VR with algorithms that removed up to 99% of tremors from PD subjects’ digital hands (76% average reduction). These data suggest PD subjects can enter VR and be asymptomatic of PD IHT. To test whether our algorithms have measurable practical utility, we created a VR painting application, FingerPaint, and tested it against TiltBrush the leading VR painting application. We empirically demonstrate that complex freeform art created in FingerPaint had up to 51% reduction in tremor-associated parkinsonian drawing irregularities (24% average reduction). Lastly, we generated a technical framework wherein movements in the real-world can be measured side-by-side with those in VR. With this framework, we observed real life parkinsonian tremors were significantly reduced by up to 87% in real life when our algorithms abolished digital hand tremors in VR (35% average reduction). This reduction was observed in 78% of subjects and was progressive as subjects had increasingly reduced real life tremors the longer they were in VR.

## Introduction

Parkinson’s disease (PD) is a neurodegenerative disease affecting approximately 6-7 million patients worldwide including one million Americans^1,2^. The disease is mainly characterized by dopaminergic neuronal loss in the in the substantia nigra that results in peripheral motor symptoms. In particular, most PD patients suffer 4-7Hz involuntary hand tremors (IHT)^3^ causing significant disability in activities of daily living (ADL)^4^. Currently, there is no cure for PD. However, several treatments are available to PD patients. These include pharmacological dopamine replacement therapies which counter dopamine depletion associated with nigral neuronal cell loss^5^. Another treatment is Deep Brain Stimulation (DBS), a type of brain surgery in which electrodes are implanted into regions containing neurons thought to be the source of tremors such as the subthalamic nucleus (STN) near the thalamus. Some STN neurons oscillate at 4-7Hz (similar to hand tremors)^6^ or at higher frequencies. DBS acts therapeutically by suppressing these signaling patterns with chronic pulses of electrical energy to these neurons^7^. Pharmacological and surgical treatments are effective, but temporary and carry a significant risk of physiological and cognitive complications including dyskinesia, dystonia, depression, and brain hemorrhage^8-11^.

Digital medicine is a field focused on using software such as video games for non-invasive clinical therapy. For instance, the experience NeuroRacer was demonstrated to improve cognitive control^12^. Recently, the FDA approved several digital medicine products for clinical use^13^ so clinicians may provide software to patients as prescriptions. Virtual Reality (VR) is digital technology where the user visual field is fully replaced with a three-dimensional digital representation. The malleability of this representation offers a significant opportunity for understanding and treating Parkinson’s disease. For instance, PD patients had improved gait and suffered fewer falls after being trained in VR obstacle courses^14^. Unfortunately, most research has focused on gait and balance^15^ with no published work to our knowledge on tremors, a cardinal symptom of PD.

We thus set out to develop a technology to enable PD patients to leapfrog the real life challenges of living with IHT by entering VR worlds in which they are tremor-free and can function at new heights in these digital metaverses. We also sought to determine if a therapeutic benefit of reduced tremors in real life can occur because the visual input provided by VR to the central circuits causing tremors may induce modified tremor-free neural signaling.

PD is thought to be a synchronization disease where groups of neurons progressively enter into pathological synchronous patterns of firings^7,16-19^. This theory predicts that interventions that break pathological synchrony (referred here as sync-reset signals) will reduce IHT. For instance, closed-loop and coordinated-reset DBS, in which a bolus of electrical energy intended to cause a sync-reset is pulsed into thalamic nuclei, reduced coherence of oscillating neurons and had a therapeutic effect on tremors^20-22^. Recently, coordinated-reset vibrotactile finger movement reduced IHT in PD patients^23^. Here the fingers of study participants were subjected to brief high-frequency trains of vibrations to deliver a mechanosensory sync-reset signal putatively to the STN, potentially inducing a sync-reset in a manner similar to STN electrical stimulation^22^. Note, the STN is a central region studied in PD, but is only one of several areas such as the ventral thalamus and pallidal tissues thought to contribute to PD. For brevity, we use the STN here as a collective placeholder of regions implicated in PD.

The thalamus is an approximately 6 cm region of the diencephalon forebrain that is composed of multiple nuclei each specializing in the input and output information of various senses and motor modalities. As such, the thalamus is where eyesight and the motor movements thought to cause tremors converge. The thalamic connection to the visual system occurs via its lateral geniculate nucleus (LGN)^24^ which is innervated directly by the optic tract. The various nuclei of the thalamus contribute to thalamic oscillations. Though these nuclei have specializations, their oscillations can be influenced by the activity of nearby nuclei. For instance, oscillations of non-visual nuclei in the thalamus can be reset by visual input from the LGN^25,26^. As such, eyesight may provide an affirmatory signal to non-visual nuclei. For example, if the eyes see real life hands tremoring at 4-7Hz, the LGN may provide oscillatory reinforcement to the STN to continue sending tremor signals to the hands at a similar frequency in a maladaptive biofeedback loop (Fig. 1).

**Fig. 1.**
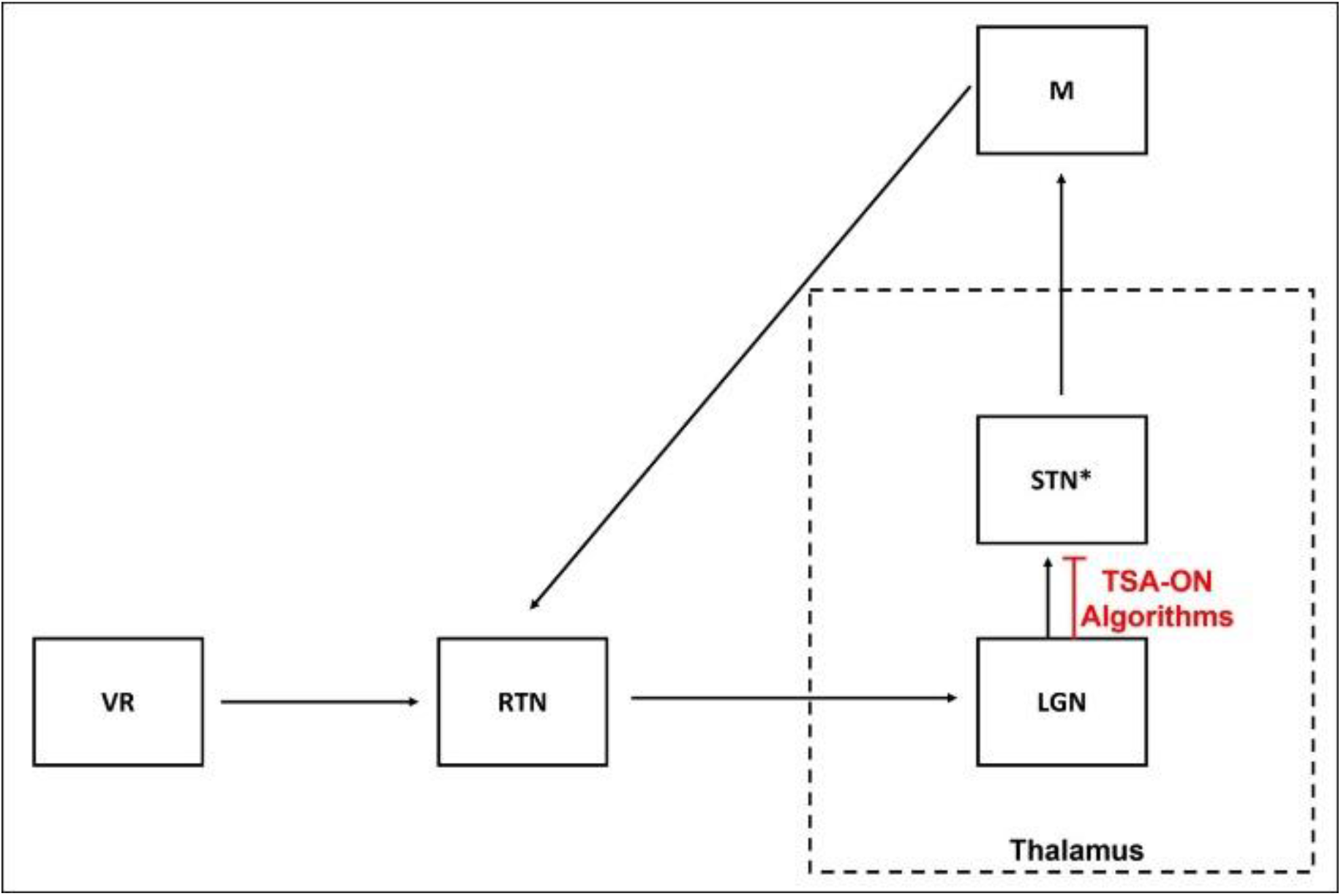
Proposed model for real life tremor reduction. The thalamus, a ^∼^6 cm sized nuclei in the diencephalon forebrain contains subnuclei that are convergence points for multiple efferent and afferent modalities. These include the lateral genicular nuclei (LGN) which receives visual input from the retina (RTN), and the subthalamic nuclei (STN) which in PD sends pathological motor (M) signals to the periphery causing patients hands to tremor. Network oscillations cascading through the thalamus are constituted by the activity of such individual nuclei, with various nuclei influencing each other. What the eyes see and outgoing tremor-causing thalamic motor efferent signals are typically in synchrony, forming what we propose to be a maladaptive pacemaker that re-enforces itself by the eyes continually confirming that the hands are shaking. As eyesight-based biofeedback is powering this pacemaker, this loop may be interrupted with Virtual Reality (VR) with Tremor Stabilization Algorithms (TSA-ON) as this intervention provides a visual signal that is out-of-sync with the STN. With the biofeedback interrupted, the STN may reset to a healthier oscillatory pattern carrying reduced tremor signals. *Note, the STN is a central region studied in PD, but is only one of several areas such as the ventral thalamus and pallidal tissues thought to contribute to PD. For brevity, we use the STN here as a collective placeholder of regions implicated in PD.

While there is no published evidence to our knowledge of such a LGN-STN interaction, it is worth considering how such a loop could be interrupted. We hypothesized that observing hands in VR (i.e., digital hands) that are not tremoring as these will move at a different phase, amplitude, or frequency than real life tremoring hands and the causative tremor oscillator in the brain. More specifically, digital hands that are not tremoring have movement at less than 1Hz thus contrasting the 4-7Hz of tremoring hands in real life and the oscillations of the STN. This desynchrony may be sufficient to provide a therapeutic sync-reset signal.

To test this hypothesis, we leveraged the fact that VR controllers are high-precision instruments that can measure position in space over time with significant sensitivity. We developed algorithms that intervene in the data stream from the controllers to their projection as digital hands in VR headsets. Briefly, controllers send positional information of the centroid space they occupy, thus giving the CPU in VR headsets their position in space including whatever is holding them, vis-à-vis real life hands. The projection of the controller into the headset is a separate process. Therefore, one could intervene or hack the projection using algorithmically treated derivatives of the controller position. Our algorithms modify the controller position by instilling a brief lagging stream of the controller position as the real time positional data. This slows the time of controller position change resulting in an illusion of digital hands with no tremors and move one-to-one with real hands in real time.

We describe in the following study the deployment of these algorithms to generate VR spaces where PD patients can be tremor-free and investigate whether tremors were reduced in real life.

## Results

In the following we describe our study design and findings. We first determine whether the Oculus VR technology can record parkinsonian tremors. We then ask whether we can algorithmically reduce tremors in VR. And investigate whether tremors were reduced in real life. We conclude by describing a new program we’ve developed to demonstrate the practical utility of reduced tremors in VR.

### Recording of parkinsonian tremors with the Oculus VR technology

The Oculus Rift primarily uses accelerometers and gyroscopes embedded within its Touch controllers to determine physical position in space. These record changes of rotational angles α, β, γ, and Cartesian axes X, Y, Z at up to 1000Hz. However, we recorded at 4Hz which is in the range of frequencies PD tremors occur at^3^, (also see Methods section). We recruited nine volunteer subjects for observation of PD associated IHT (no inclusion/exclusion criteria other than self-report of PD, Table 1). PD subjects were placed in a virtual kitchen where they could perform tasks such as placing bread into a toaster and pouring cereal into a bowl. This familiarized the PD subjects with the Oculus headset and its Touch controllers. We then loaded a stock virtual testing environment and instructed subjects to perform a postural tremor test (PTT) in which the subjects stabilize their backs against a wall and position their hands in the air against gravity for two minutes (Fig. 2). This position places the subjects’ hands in a posture such that they must maintain gait against gravity and is known to elicit postural parkinsonian tremors^27,28^.

**Table 1.**
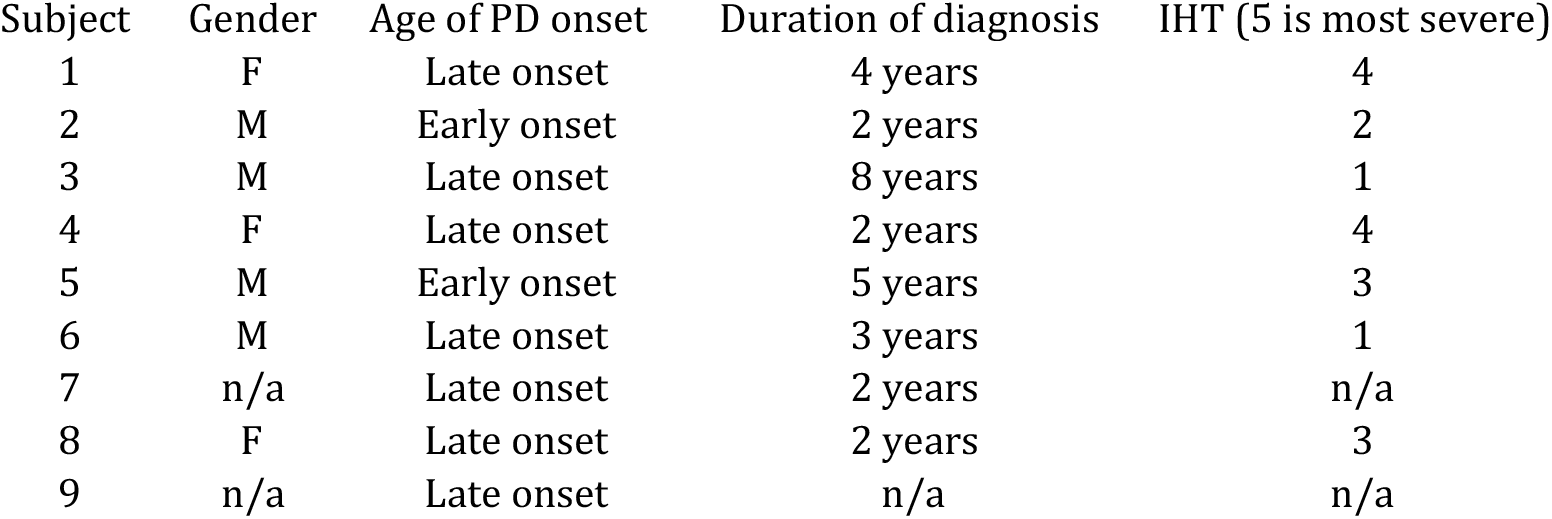
Demographic information of study participants. Note patients were not screened for prognosis and severity of PD, or whether on oral or surgical medication (i.e., there were no inclusion/exclusion criteria other than PD self-report). Early onset defined by diagnoses less than 50 years. Severity of involuntary hand tremor (IHT) scale ranges from 1 (no tremors) to 5 (severe tremors). All data was self-reported.

| Subject | Gender | Age of PD onset | Duration of diagnosis | IHT (5 is most severe) |
| --- | --- | --- | --- | --- |
| 1 | F | Late onset | 4 years | 4 |
| 2 | M | Early onset | 2 years | 2 |
| 3 | M | Late onset | 8 years | 1 |
| 4 | F | Late onset | 2 years | 4 |
| 5 | M | Early onset | 5 years | 3 |
| 6 | M | Late onset | 3 years | 1 |
| 7 | n/a | Late onset | 2 years | n/a |
| 8 | F | Late onset | 2 years | 3 |
| 9 | n/a | Late onset | n/a | n/a |

**Fig. 2.**
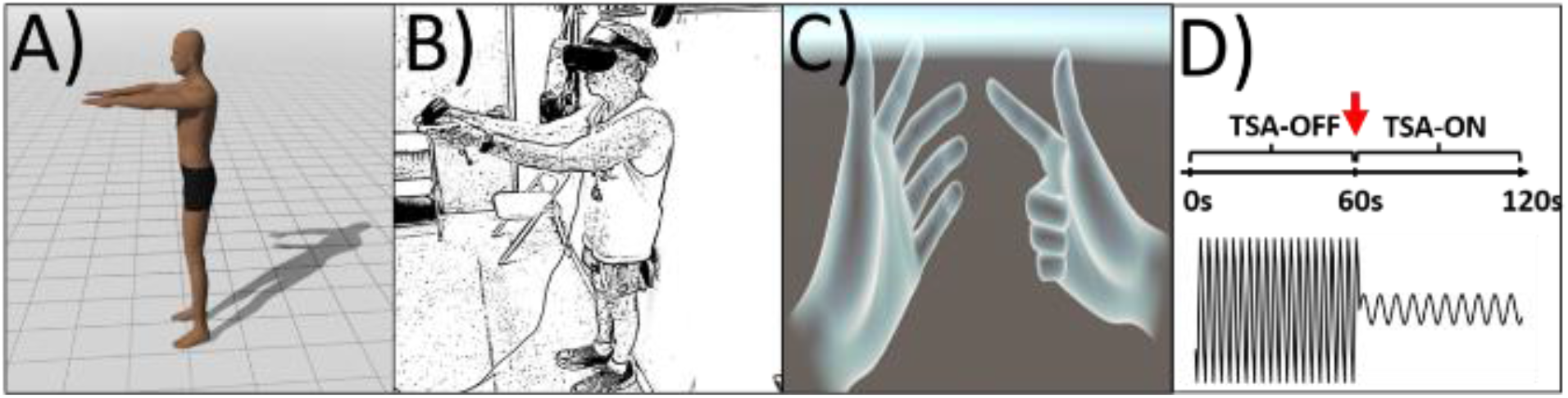
The postural tremor test (PTT). Subjects performed a PTT by positioning their arms at 90 degrees relative to their body (**a**). The upper body posture was maintained against gravity for two minutes to elicit parkinsonian tremor. Subjects stabilized their backs against a wall for comfort and for minimization of non-specific body movements becoming reflected in hand tremor measurements (**b**). Representative image depicting what subjects observed in VR: the VR space was sparse as it consisted only of a stock Unity software environment with a floor and endless blue horizon, and a digital depiction of their hands in VR. Image is from the Oculus Avatar SDK ver 1.0 (**c**). In the PTT (**d**), Tremor Stabilization Algorithms were off during the first 60 seconds (TSA-OFF) and then turned on in the second 60 seconds (red arrow, TSA-ON). Recording was continuous during the two segments. Mock trace at bottom shows expected record of Oculus Touch controller movement measuring tremors during the PTT.

During the PTT, Tremor Stabilization Algorithms (TSA) were either turned off or on (TSA-OFF, TSA-ON). Specifically, the first minute of the PTT was TSA-OFF and the second minute was TSA-ON (Fig. 2d). We found that the rotational signature of IHT was most discernible with tremor-associated changes in hand rotation (Supplemental Fig. 1a-c) compared to changes in hand position (Supplemental Fig. 1d-f). Similar work using different instrumentation also found rotation to be a reliable measure of PD tremors^29^. Importantly, we recorded the positional information of PD subjects’ hands in the real-world (i.e., the position of the Oculus Touch controllers in physical space) simultaneously with the virtual position of the subjects’ digital hands in VR. This enabled segregation of effects on IHT in the real-world from those in VR. As describing results in physical space and virtual space can easily become confusing: we define data of hand positions in physical space as classical reality (CR, green traces in Supplemental Fig. 1 and Fig. 3) and data of digital hand positions in virtual space as virtual reality (VR, yellow traces in Fig. 3).

**Fig. 3.**
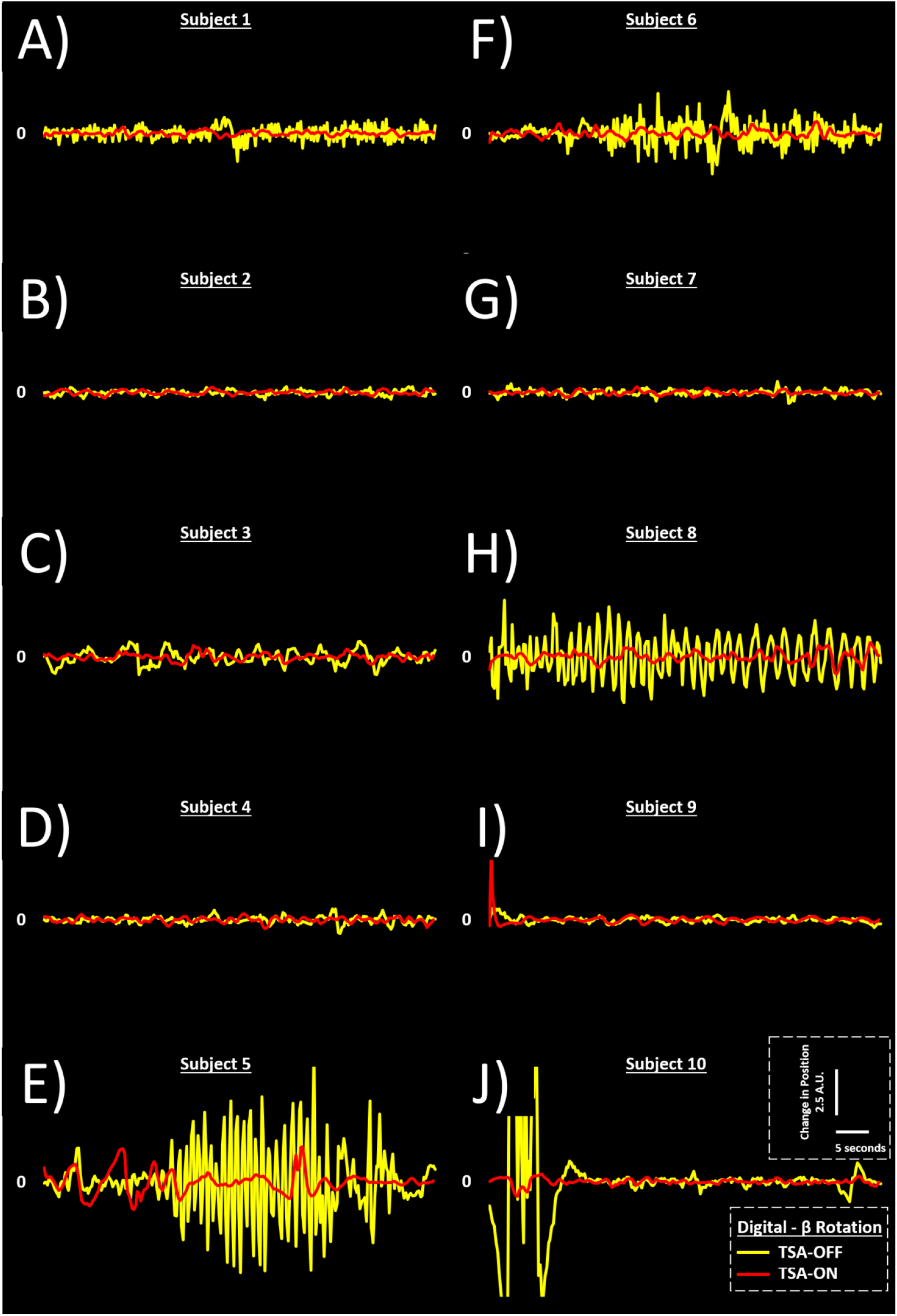
Parkinsonian involuntary hand tremors can be abolished in VR. Representative β rotational changes of digital hand in VR are presented here. TSA were either turned off (TSA-OFF, yellow) or on (TSA-ON, red) for all PD subjects (AI) and control subject(J). Several subjects (see **a, c, e, f**, and **h**) had IHT qualitatively reduced in VR with TSA-ON compared to TSA-OFF. Subjects did not undergo screening (see Table 1) thus variability in IHT was expected. High amplitude spikes in control subject trace (**j**) were likely due to whole body movement and did not have high frequency character of Parkinsonian IHT.

VR data of digital hands from subjects captured with TSA-OFF showed IHT with high-frequency oscillations (Fig. 3a, c, e, f, and h). These oscillations were qualitatively reduced with TSA-ON in VR. The reductions were instantaneous and persistent indicating robust temporal sensitivity and little algorithmic habituation. Data from a control subject who did not have PD showed no sign of IHT in VR (Fig. 3j) and CR (data not shown). Together, these data demonstrate the Oculus Rift as a capable recorder of IHT, that hand rotation caused by tremors provides a robust IHT signature, and that qualitative preliminary inspection suggests that our algorithms reduce IHT of digital hands in VR.

### Reduction of tremors in VR by our algorithms

To quantitatively determine if our algorithms reduced tremors in VR, we applied continuous wavelet transformation to our tremor recordings. The frequency content of oscillating signals is described completely in a lossless manner by convoluting it with appropriate wavelets and examining the variance of the scale-by-scale decomposition^30,31^. Application of the Morlet wavelet (*morl*) on VR tremor recordings (β rotation of tremor dominant hand; representative raw trace in Fig. 4a) resulted in scalograms of the decomposed signal for TSA-OFF (Fig. 4b) and TSA-ON (Fig. 4c). The scalograms carry data of time-associated higher frequency features at low scales and lower frequency features at high scales. High coefficient magnitudes in the scalogram indicate higher power events^31^. Thus the totality of the scalogram both in terms of frequency space and coefficient magnitude amounts to variation that describe the severity of PD tremors (Fig. 4d). We quantified variance of scalograms for all PD subjects (n = 9) to assay for severity. The mean of the variance of each subject for TSA-ON and TSA-OFF indicated all the digital hands of all subjects (100%) had reduced tremors (75.5% average reduction, p = 0.01, min reduction = 48.6%, max reduction = 98.7%). Subjects were grouped by age of PD onset and duration of diagnosis; no clear patterns were observed (Fig. 4e and f). To gain insight on the temporal characteristic of tremor reductions, the mean variance of TSA-OFF and TSA-ON across all subjects (Fig. 4g) was segmented to four 15s bins. We found that in all time bins, VR tremors were reduced or nearly eliminated (Fig. 4h). Tremors were significantly reduced by 40% for 0-15s, by 71% for 15-30s, by 75% for 30-45s, and finally by 97% for 45-60s (p *<* 0.05 for time bins within 15-60s). The percentage-wise trend of these reductions indicate a ceiling of tremor reduction was approached where no further tremor reduction would be possible (gray line in Fig. 4h). This suggests that our algorithms saturated possible gains in VR tremor reduction, thus highlighting their effectiveness. The data was qualitatively supported by PD subjects self-reports of not seeing their digital hands tremor. It is worth noting TSA-OFF tremor severity was markedly lower during in the 0-15s bin thus the lower extent of tremor reduction compared to other bins was likely because there were fewer tremors at baseline. Overall the severity of tremors we observed was varied in time which is a well-characterized feature of PD^32^. Some postural tremors are known to have a ^∼^10s latency (i.e., reemergence^33^) which could explain the 0-15s bin data.

**Fig. 4.**
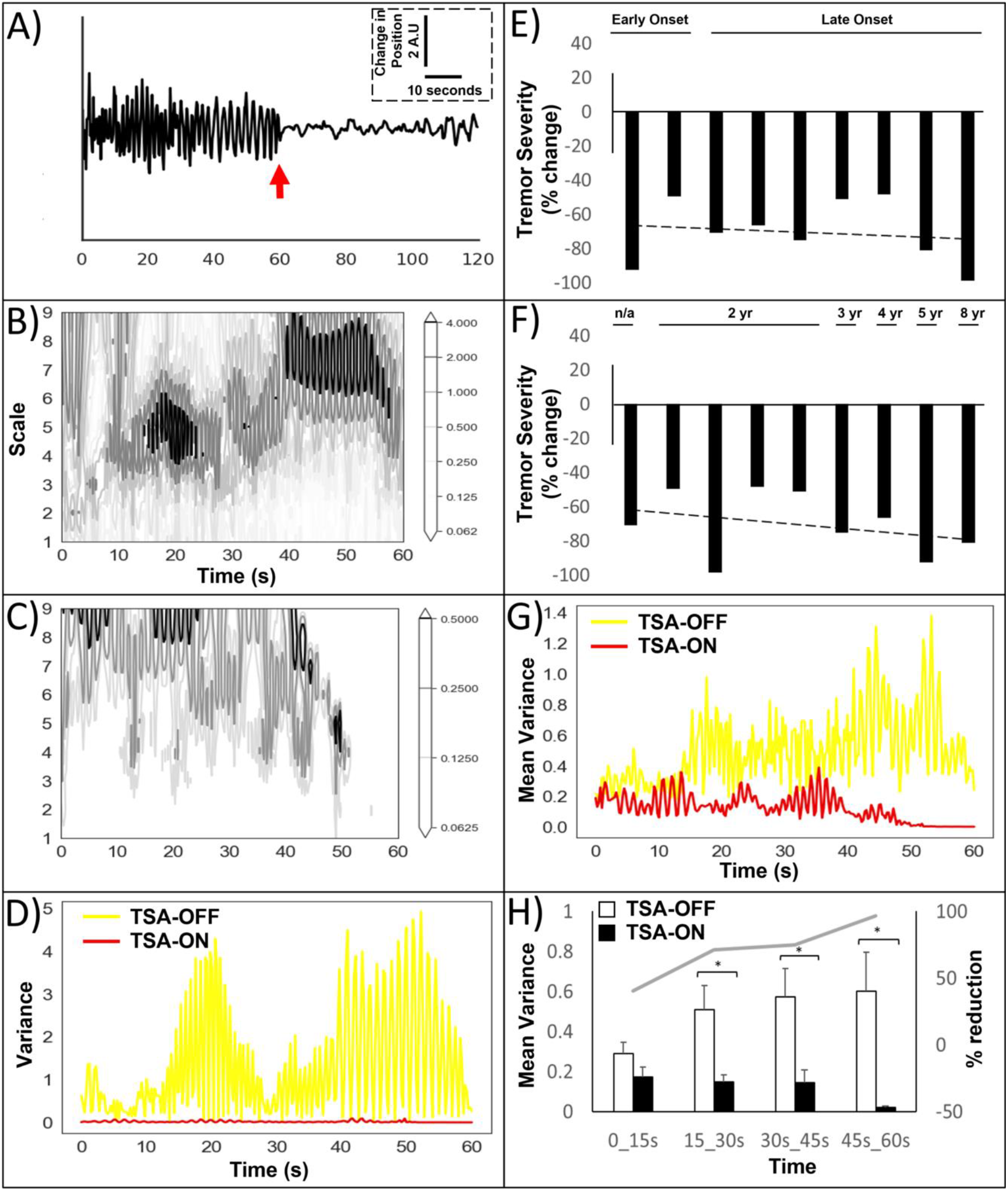
Wavelet decompositions show near complete cessation of tremors in Virtual Reality during TSA-ON. Representative full trace β rotation of a subject tremor dominant hand in VR during the PTT (**a**). Red arrow in (**a**) indicates start of TSA-ON. The frequency content of the trace is described completely in a lossless manner by convoluting it with a wavelet and examining the variance of the scale-by-scale decomposition^30,31^. The Morlet wavelet (*morl*) was applied to the first 60s and second 60s segments of traces which correspond to TSA-ON, TSA-OFF, respectively. Scalograms of wavelet coefficients in (**a**) are plotted for TSA-OFF (**b**) and TSA-ON (**c**). Magnitude of coefficients in the scalogram is represented in the accompanying colorbars. Increased magnitude and complexity of the contours (colorbar scales are unequal) indicate increased variance. The variance of these scalograms is displayed in (**d**) and indicate near complete cessation of tremors with TSA-ON. The relative change in tremor severity (TSA-OFF/TSA-ON mean variance) with linear trendline (dashed line) of individual subjects (**e** and **f**) indicated all subjects had reduced tremors. These are arranged by age of onset where less than 50 years is considered early onset (**e**) and duration of time from initial diagnoses of PD (**f**). The mean of variances of all PD subjects (n=9) is plotted in (**g**) and were binned into 15s segments to gain insight on the temporal characteristics of tremor reduction (**h**). Statistical analyses. *, p*<*0.05. Gray line in (**h**) is percentage reduction in tremors derived from mean variances.

### Reduction of tremors in CR (i.e., real life) by our algorithms

As noted above, we captured IHT signatures separately in VR and CR thus providing an ideal condition with which to test if reduced IHT in VR results in reduced IHT in CR. As expected, data of CR with TSA-ON retained high-frequency oscillations in most PD subjects (representative PD subject in Supplemental Fig. 2a). However, CR IHT was notably reduced in one PD subject (Supplemental Fig. 2b) suggesting other subjects exhibited reduced real-world tremors that were perhaps undiscernible in raw frequency traces. To address this, we applied wavelet transformation to decompose the CR data (Fig. 5) in a manner similar to VR data described above. The mean variance of these transformations for all subjects (n=9) indicated that real-world tremors were reduced by 35.3% (p = 0.02, min reduction = 28.6% increase, max reduction = 88.7%). Segregating this data further, seven of nine subjects (77.7%) had real-world tremors that were reduced by more than 10% (representative subject in Fig. 5a-d). No clear patterns were observed for age of PD onset or duration of diagnosis (Fig. 5e and f). To gain greater insight on the temporal characteristic of tremor reductions, the mean variance of CR TSA-OFF and TSA-ON was segmented to four 15s bins as done above for VR data. The mean variance of scalograms of all PD subjects (n = 9, Fig. 5g) indicated tremors were increased by 32% in the first 15s bin (Fig. 5h). However CR tremors diminished by 13% for 0-15s, by 22% for 15-30s, by 33% for 30-45s, and finally by 77% for 45-60s (p *<* 0.05 for 30-45s time bin, Fig. 5h). The percentage-wise trend of these reductions indicate CR tremors were progressively reduced with time (gray line in Fig. 5h). Further experimentation of time and algorithmic design may yield greater reductions of CR tremors.

**Fig. 5.**
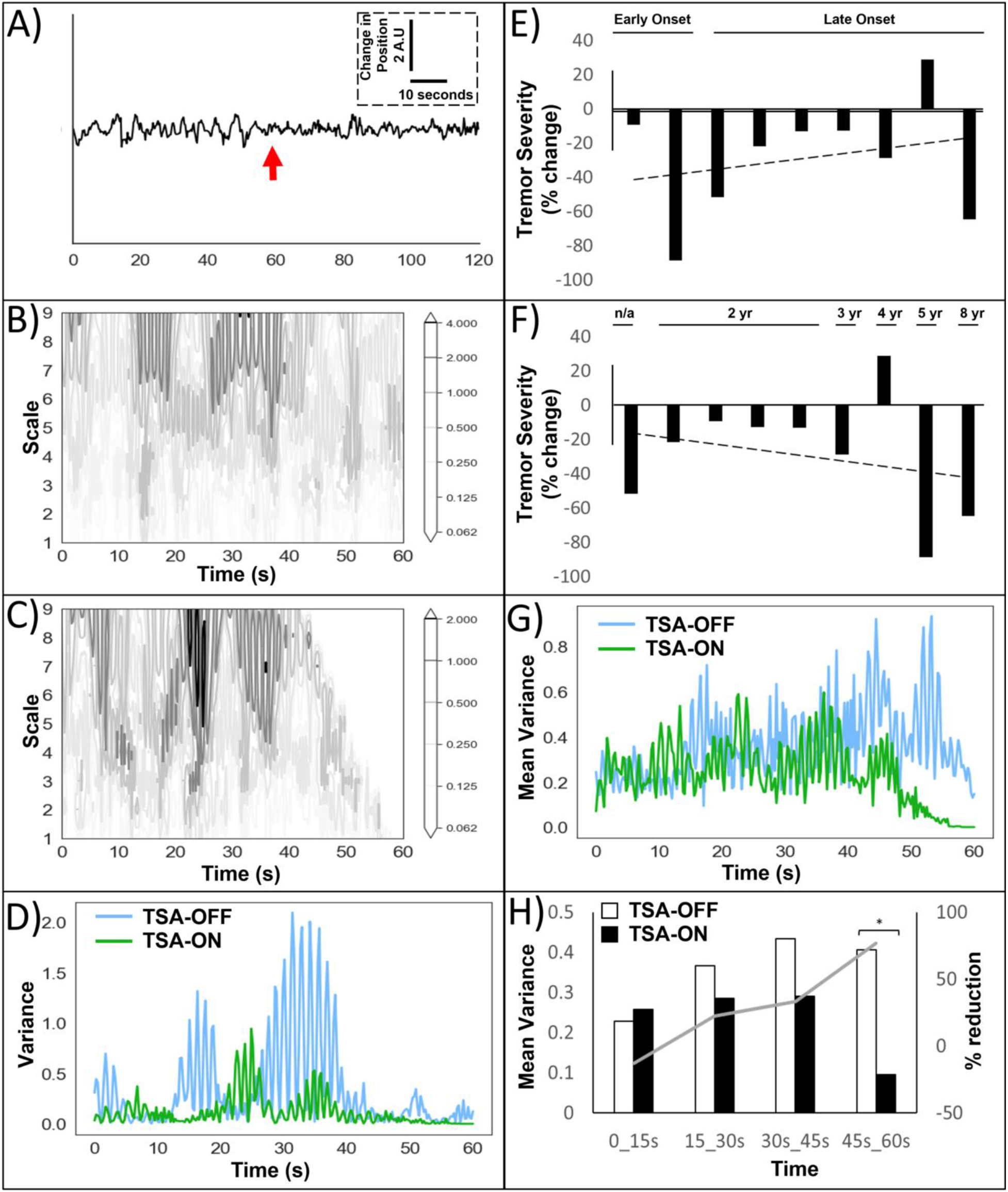
Wavelet decompositions show reduction of tremors in Classical Reality (i.e., real world) during TSA-ON. Representative full trace of β rotation of a subject tremor dominant hand in CR during the PTT (**a**). Red arrow in (**a**) indicates start of TSA-ON. The Morlet wavelet (*morl*) was applied to the first 60s and second 60s segments of traces which correspond to TSA-ON, TSA-OFF, respectively. Scalograms of wavelet coefficients in (**a**) are plotted for TSA-OFF (**b**) and TSA-ON (**c**) with the magnitude of coefficients in the scalogram represented in the accompanying colorbars. Increased magnitude and complexity of the contours (colorbar scales are unequal) indicate increased variance. The variance of these scalograms is displayed in (**d**) and indicate a notable reduction of tremors with TSA-ON. The relative change in tremor severity (TSA-OFF/TSA-ON mean variance) of individual subjects indicated seven of nine subjects (77.7%) had greater than 10% reduction of tremors (**e** and **f**). Subjects are arranged by age of onset where less than 50 years is considered early onset (**e**) and duration of time from initial diagnoses of PD (**f**). The mean of variances of all PD subjects (n=9) is plotted in (**g**) and were binned into 15s segments to gain insight on the temporal characteristics of tremor reduction (**h**). Statistical analyses: *, p*<*0.05. Trendline in (E-F) is linear best fit. Gray line in (**h**) is percentage reduction in tremors derived from mean variances.

### FingerPaint, a new program to demonstrate practical utility of our framework and algorithm

PD patients in professions requiring fine motor skills oftentimes become limited in their work and face early retirement because of difficulties with tremors. For instance, graphic designers and artists require precise articulation of finger motions to generate their works. Amongst the most popular and innovative applications to create art that is available for the Oculus platform is TiltBrush (Google, Menlo Park, CA). TiltBrush features a myriad of paintbrushes allowing for complex artwork enjoyed by casual users and increasingly applied in professional settings.

However neither TiltBrush nor any other VR programs offer stabilization of tremors thus reducing its utility for PD patients. To demonstrate the practical utility of our framework and algorithms, we constructed an application for creating paintings in VR. The program, called FingerPaint features TSA-ON algorithms and allowed users to employ several paintbrushes to generate three-dimensional paintings in VR. A PD subject created a series of simple drawings (i.e., circles, triangles, and squares) as well as a complex drawing of a face in TiltBrush and FingerPaint for side-by-side comparisons (Fig. 6). In the simple drawings on TiltBrush, irregularities in articulation were evident where brush strokes transitioned in direction such as at the corners of triangles and squares (henceforth referred to as parkinsonian drawing irregularities). Similar regions in FingerPaint instead featured smooth transitions (see insets and arrows in Fig. 6a and b). These differences were also found in complex drawings of faces (Fig. 6c and d). The parkinsonian drawing irregularities that generally were isolated in the simple drawings in Tiltbrush were compounded with the greater complexity the face drawing demanded and resulted in a visually distinct impression in TiltBrush compared to FingerPaint. For instance, strings of head hair and eyebrows were squiggly and inconsistent primarily in Tiltbrush.

**Fig. 6.**
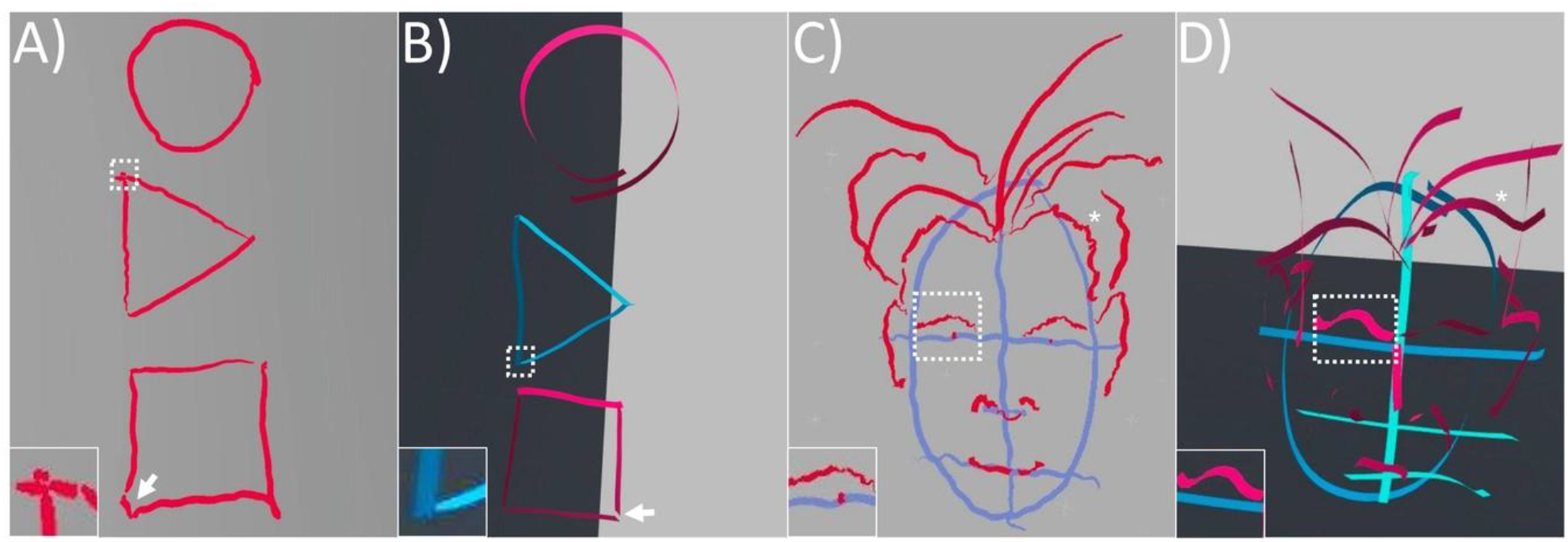
Comparison of fine-motor skills in VR environments with and without Tremor Stabilization Algorithms. We created an application for painting in VR with brush strokes powered by our tremor stabilization algorithms. This program, called FingerPaint was compared to Tiltbrush (Google, Inc), the leading application for painting in VR. Drawing of simple shapes (**a** and **b**) revealed parkinsonian fine motor irregularities in TiltBrush (**a**) but not in FingerPaint (**b**). These irregularities were particularly notable in brush direction transition zones such as the corners of triangles and boxes. See insets and arrows in a-b respectively. The drawing of more complex objects such as faces revealed greater and more compounded irregularities (**c** and **d**). For instance, the drawing of subtle features such as eyebrows and hair strings were irregular in TiltBrush (**c**) and smooth in FingerPaint (**d**). See insets for zoom-in of eyebrows and asterisk for hair strings in **c** & **d**.

Current methods of quantifying these parkinsonian drawing irregularities include drawing clocks or Archimedes spirals^34,35^. These methods are not empiric as they depend on human judgement which could vary from test to test. We hypothesized that drawings with and without parkinsonian tremor were primarily differentiated by micro-curvatures in the local structure of drawn lines and thus tools that measure such local structure would be able to empirically quantify parkinsonian drawing irregularities. We thus sought programs that measure angular curvature and discovered FilFinder, a program designed to study cosmic dust emission and star formation^36^. FilFinder traces and adaptively decomposes complex structures down to one-pixel width filaments and is thus ideal for isolating complex features such as hair in drawings.

Moreover, FilFinder applies the recently developed Rolling Hough Transform (RHT)^37^ on filaments to quantify the distribution of angles in micro-curvatures. We validated FilFinder on this capacity by generating lines which we introduced salt and pepper noise to artificially simulate tremors (Supplemental Fig. 3). We found that the distribution of the angles in micro-curvatures reported by FilFinder RHT increased proportionately to the amount of simulated tremor. Depiction of these angles in 2_θ_ radar charts revealed a cluster which became increasingly large proportionally to the simulated tremors (Supplemental Fig. 4a-d). In particular, each doubling of simulated tremors resulted in an average increase of approximately 17% in the distribution of micro-curvature angles (Supplemental Fig. 4e). Together these data validate that FilFinder with RHT is robustly sensitive in a dose dependent way to the local structure of micro-curvatures caused by simulated tremors and is therefore suitable to empirically quantify parkinsonian micro-curvatures featured in complex artwork.

We thus applied FilFinder to FingerPaint and TiltBrush face drawings (Fig. 7a, b, f, and g). FilFinder isolated multiple continous one-pixel width filaments for each drawing (n = 6 TiltBrush; n = 5 FingerPaint). These filaments were of similar average length (380.3 pixels TiltBrush; 396.42 pixels FingerPaint). RHT analyses as described above of these filaments determined that FingerPaint had on average 24% less micro-curvatures compared to TiltBrush (p = 0.02, Fig. 7l). Comparison of an individual filament of the same feature (strand of hair in forehead of the faces and extending to right side of the faces – Fig. 7c, d, h, and i) indicated that FingerPaint had 50.9% less micro-curvatures of this feature compared to TiltBrush (Fig. 7e, j, and k). This demonstrated that TSA-ON algorithms in FingerPaint reduced the parkinsonian drawing irregularities in a complex freeform work of art suggesting our algorithms facilitated a partial restoration of the subjects’ capacity to perform a fine-motor skilled activity.

**Fig. 7.**
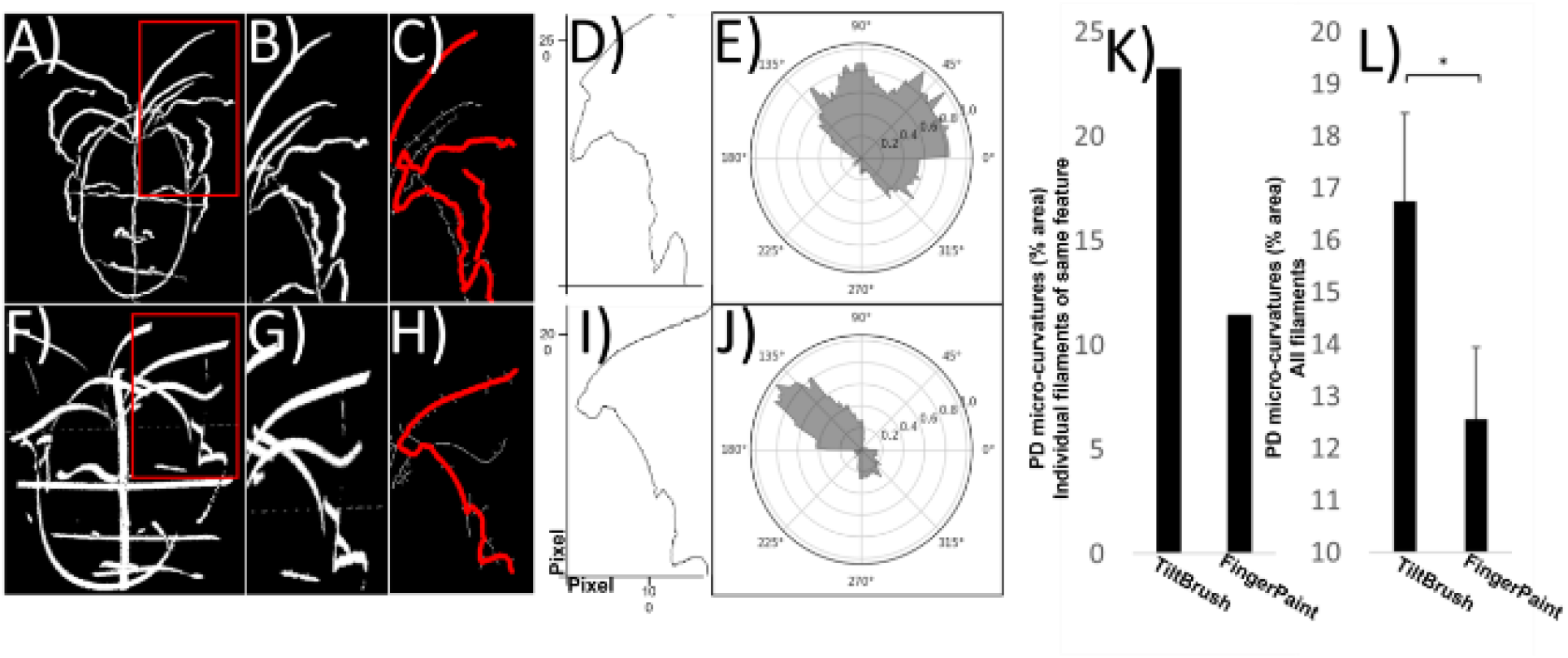
Use of FingerPaint shows reduction in parkinsonian tremor irregularities. Paintings from Fig. 6 were binarized (**a** and **f**). A section of binarized image containing complex features such as hair (red box in **a** and **b, f** and **g**) was tracked by FilFinder (red overlay in **c** and **h**). A one-pixel wide filament of continous features were generated from these traces (representative traces in **d** and **i**). Rolling Hough Transform (RTH) was performed on the filaments to quantify micro-curvatures due to tremors. RTH coefficients are plotted as angular distributions in the 2_θ_ chart (filaments **d** and **i** represented in **e** and **j**). Comparison of these filaments which represent the same complex feature (strand of hair in forehead of the faces and extending to right side of the faces (**b, g, d**, and **i**) indicated that FingerPaint had 50.9% less micro-curvatures compared to TiltBrush (**k**). The area of these distributions for all filaments indicated that micro-curvatures were reduced on average by 25% in FingerPaint relative to TiltBrush (**l**). n = 6 filaments for TiltBrush, n = 5 filaments for FingerPaint. Average length of filaments was 380.1 pixels for TiltBrush and 396.4 pixels for FingerPaint.

## Discussion

Multidisciplinary neurotechnology holds the promise of understanding and non-invasively treating neurodegenerative diseases. In this study, we combined neuroscience together with the nascent field of medical virtual reality to generate a technology to enable PD patients to leapfrog the real life difficulties of living with parkinsonian IHT by entering VR metaverse worlds in which they are tremor-free and can function at new heights.

We generated algorithms for reducing digital tremors and implemented them into the Oculus Rift VR platform. This led to a series of important observations. First, we established the Oculus Rift is a potent measurement device for parkinsonian IHT. Secondly, we determined parkinsonian tremors can be abolished in VR with algorithms that eliminate tremors from digital hands in 100% of subjects. Thirdly, we developed the app FingerPaint which is powered by our tremor reduction algorithms and demonstrated that simple and complex artwork is more articulated and had 24% less parkinsonian drawing irregularities compared to the industry leading painting app Tiltbrush. Lastly, we employed wavelet decomposition of tremors and determined that 78% of PD subjects had their real life tremors reduced by up to 89% when our tremor reduction algorithms were on. This reduction was progressive as subjects had increasingly reduced tremors the longer they were in VR.

Our study design focused on postural tremors as these are well documented and are amenable to an active standing up position which is customary for interacting with VR worlds (see Methods). Postural tremors in the context of PD were thought to be an extension of resting tremors^38^. However it is increasingly clear postural tremors are a heterogeneous group of tremors. For instance, reemergent postural tremors have a ^∼^10s latency as their distinguishing feature (see Supplemental Fig. 2b)^33^. More recently, a novel class of postural tremors was described by Dirkx *et al*.^28^. These had no latency, had a higher frequency than resting and reemergent tremors, and were dopamine insensitive. Our data undoubtedly contains a diversity of postural tremor subtypes particularly because our open recruitment study design (no inclusion/exclusion criteria other than self-report of PD) yielded a heterogeneous cohort of subjects. Further discernment of postural tremor subtypes will inform us to which tremors are most responsive to stabilization in virtual worlds and real life.

Notably, our algorithms abolished nearly all tremors in all subjects in VR. The practical utility of this achievement cannot be understated. There are many essential tasks that PD patients can perform in virtual reality for real life impact. To illustrate this, we focused on a group of PD patients whose professions require fine motor skills as they oftentimes face early retirement related to difficulties with tremors. These include graphic designers and artists. We thus created the app FingerPaint powered by our TSA-ON algorithms. Compared to the leading VR painting app TiltBrush, FingerPaint reduced parkinsonian drawing irregularities and restored articulate drawings of simple and complex paintings. The irregularities we observed in TiltBrush included stoppages at transition points of brush strokes. For instance when turning a corner while drawing a triangle. Such stoppages are likely due to rigidity, a cardinal parkinsonian symptom. We observed several other irregularities and their totality resulted in complex drawings that were notably modified in affect in TiltBrush compared to FingerPaint. For instance, eyebrows were wavy and twisted in TiltBrush while smooth and life-like in FingerPaint. These irregularities were noticeable by eye but are hard to empirically quantify.

We hypothesized that drawings with and without parkinsonian tremor were primarily differentiated by micro-curvatures in the local structure of drawn lines and thus tools that measure such local structure would be able to empirically quantify parkinsonian drawing irregularities. Towards this, we repurposed FilFinder, a tool typically used by astronomers to study cosmic dust clouds and the formation of new stars to extract micro-curvature features in simulated parkinsonian tremors, and Rolling Hough Transform to present the distribution of angles of the micro-curvatures in the simulated parkinsonian tremors. Application of this methodology to complex free-form drawings performed in FingerPaint and TiltBrush demonstrated that parkinsonian drawing irregularities were reduced by up to ^∼^51% in FingerPaint. Together this work demonstrates fine motor skills that are lost due to PD tremors can be restored in VR.

Currently, there is no cure for PD. Available drug and brain surgery treatments are invasive, temporary, and carry a significant risk of physiological and cognitive side-effects including dyskinesia, dystonia, depression, and brain hemorrhage^8-11^. These treatments are thought to suppress pathological synchronization of neurons^7^. In particular, it is thought that sync-reset signals that break pathological synchrony will reduce IHT^7,16-19^. This is supported by the observation that pulsing electricity into pallidal or thalamic nuclei reduced coherence of oscillating neurons and had a therapeutic effect on tremors^20-22^. However, the influence of other sensory modalities in reducing PD tremors is not well characterized. A recent work by Syrkin-Nikolau and colleagues showed that a sync-reset signal delivered peripherally through the mechanosensory system can reduce tremors^23^. We extended this thinking by asking if a similar therapeutic effect occurs with visual sensory pathways and hypothesized that virtual reality spaces where PD patients’ digital hands are not tremoring would create a visual representation that is out of sync with the thalamic firing that is causing the tremors in real life. The out of sync visual representation would thus facilitate desynchrony in the thalamus and non-invasively reduce tremors (Fig. 1). Our results support this reasoning as we observed reduction of tremors in real life when our algorithms stabilized the digital hands of our subjects.

Unlike VR tremor reduction which practically abolished digital tremors rapidly, tremor reduction in real life progressively increased over time. This suggests more tremors were reduced the longer subjects were in VR. Further experimentation with greater time intervals is needed to fully explore this observation. Algorithmic experimentation on digital hand tremor stabilization may also yield further reduction of real life tremors.

Another question worth considering is whether tremors remained reduced when subjects left VR and returned to the real-world. To gain insight on this question, it is worth considering the carryover persistence of other externally-delivered sensory biofeedback interventions. For instance the anti-tremor therapeutic effect of coordinated-reset vibrotactile finger movement persists up to 1 month following treatment^23^. Similarly, Delayed Auditory Feedback (DAF) wherein the sound of subjects voices is delayed by hundreds of milliseconds before replay reduces stuttering up to five years following the treatment^39,40^. These observations show the potential of sensory biofeedback interventions to persist long term. Thus, an area of future study is to investigate if real life tremor reductions described here persist long-term.

In conclusion, in this study we observed that our algorithms abolished tremors from the digital hands of PD subjects in VR worlds, which was associated with significant reduction of tremors in real life. Compared to drug and surgical approaches, our approach is non-invasive digital medicine and is based on light entering the visual system to putatively restore healthy thalamic oscillations. This is a new paradigm with a sound theoretical framework to build on.

## Methods

### Study subjects

Subjects were informed to sign-up for our study via an email newsletter prepared by stoPD, Inc., a non-profit organization for PD patients (www.stoppd.org). Subject data can be found in Table 1. A control subject served primarily as a sanity check of technical pipelines. PD subjects were not screened for prognosis and severity of PD, or whether on medical or surgical treatment (i.e., there were no inclusion/exclusion criteria other than self-report of PD). Advarra (Columbia, MD) Institutional Review Board (IRB) determined this study to be exempt from IRB oversight. All data were collected with subject informed consent, anonymized during capture and analyses, and analyzed in a HIPAA-compliant Amazon Web Services (AWS, Amazon, Seattle, WA) computational pipeline. Measurements were recorded with the utmost safety measures, to ensure that our subjects had a safe experience. All methods were carried out in accordance with relevant guidelines and regulations.

### Virtual Reality hardware

We employed the Oculus Rift S headset and its Touch controllers (Facebook, Menlo Park, CA). The Oculus was tethered to a laptop running a Unity environment (Unity v2019.1.5f1, San Francisco, CA) for the capture of changes in rotation and Cartesian axes. The Oculus VR space that subjects observed was a sparse, stock Unity software environment with a floor and endless blue horizon, and a digital depiction of their hands in VR.

### Postural Tremor Test

Subjects performed the postural tremor test (PTT) by stabilizing their backs against a wall and positioning their hands in the air against gravity for two minutes (Fig. 2). This position places the subjects’ hands in a posture such that they must maintain gait against gravity and is known to elicit postural parkinsonian tremors^27,28^. We choose to focus on postural tremors, rather than resting tremors, as it is amenable to standing up with active hands, which is the common body posture for using VR. We chose two minutes as the recording period to be considerate of the fatigue subjects may feel from holding their hands up, and this period falls within the timeframe at which postural tremors occur^28,33^ at their maximum tremor severity under provocation^41^. Within the two minutes subjects were performing the PTT, Tremor Stabilization Algorithms were kept off during the first 60 seconds, and then turned on in the second 60 seconds. Recording was continuous during the two segments as recommended^29^. Head tremor was not considered in analyses because it’s impact was likely absorbed by the wall subjects leaned their backs on, and it is rare in PD^42^.

### Tremor stabilization algorithms

The tremor stabilization ideation and algorithms developed in this study are along the lines of equalizer algorithms from up to six decades ago, well before the age of modern VR^43-46^. Our implementation works by capturing the position and rotation of users CR hands at 200Hz. At any given moment in time, we employed the lagging 8% of this data stream to form a moving average centroid that positioned users’ digital hands in VR. This mimicked smooth hand motions.

### Data collection

For analyses, CR and VR hand positions were stored in data files at 4Hz. The sampling frequency for hand motion was 200Hz, yet we captured data at 4Hz. Capturing hand motion for real-time one-to-one representation of digital hands to real life hands requires extremely fast sampling frequency. However, similarly to previous work^47^, it was sufficient to record at 4Hz as this falls within the range parkinsonian tremors occur^3^, without excessively bloating data files. Parkinsonian tremors are not perfectly sinusoidal thus Nyquist sampling criteria need not apply fully.

### Data processing

Data files were uploaded to a Jupyter Notebook on an AWS EC2 server. Within the Jupyter environment, data was processed with python using Pandas, Numpy, Scikit-Learn and Scipy libraries.

We observed that digital hands in VR temporarily moved to a new position with activation of tremor stabilization algorithms. This resulted in outlier positions that were identified with *scipy*.*stat*.*zscore* (z threshold *>* 3) and removed with Pandas.

During the PTT, subjects often shifted their bodies against the wall. This introduced drift in the data that was removed with the linear leastsquares function *scipy*.*signal*.*detrend*. High frequency noise oscillations (*>*=50 Hz) were removed with *scipy*.*signal*.*butterworth*^48^. Data was centered at y-axis = 0.

TiltBrush and FingerPaint drawings were binarized with the OpenCV and Pillow libraries. These binarized images were cropped and inputted to FilFinder. We modified FilFinder with custom code to output raw data in our Jupyter environment, otherwise all settings were default. The theory and implementation of FilFinder is described by Koch and Rosolowsky^36^. The Rolling Hough Transform (RHT) is a novel machine vision algorithm used by FilFinder to detect and parameterize linear structure^37^.

### Statistical analysis

Continuous wavelet transformations were performed on PTT tremor data using the pywavelets library. We utilized the Morlet wavelet (*morl*) with 10 scales across all subjects. The absolute values of wavelet coefficients were utilized to summarize positive and negative hand movements. The variance of scalograms, the mean of variances, the means of the areas of RHT 2_θ_ radar charts, and t-tests of these means were obtained with python’s statistics library. All data was examined for normality using the Shipiro-Wilk test and manual inspection with Q-Q (Quantile-Quantile) plots. Tremoring has non-linear characteristics^49^ and thus this data was non-normal. Data was logarithmically made more parametric with sklearn.preprocessing.PowerTransformer. TSA-OFF and TSA-ON are sequential thus a paired, two-tailed Student’s t-test was employed this on data. An unpaired, two-tailed Student’s t-test was utilized to ascertain the statistical difference of micro-curvatures in drawings in TiltBrush relative to FingerPaint. p-values of p*<*0.05 were considered statistically significant. Average of variances data are presented as mean ± SEM.

## Data Availability

All data produced in the present study are available upon reasonable request to the authors.

## Data availability

Anonymized datasets generated during and/or analyzed during the current study are available from the corresponding author on reasonable request.

## Acknowledgements

Special thanks to Games for Change. To Olivia Goldman and Jared Cameron (NeuroStorm, Inc) for their helpful insight on this work, to Kurt Young (Mokuni Games, LLC) for his inspiring work expanding the potential of medical VR and to Alex Montaldo (stoPD, Inc.), Roberta Marongiu (stoPD, Weill Cornell Medicine) and Nisha L Sajnani (NYU) for coordinating subjects for this study and for exploring novel ways to tackle Parkinson’s disease. Lastly, a posthumous thank you to Jose Zambrano for being an engaging and enlightening part of our development team. This study was funded by NeuroStorm, Inc.

## Author contributions

EGN conceived and directed the experimental paradigm. JC developed and implemented the tremor stabilization algorithms with EGN. JC, AG and RC generated the virtual reality environments. RM and stoPD coordinated study subjects. EGN, JC and AG performed the experiments. DC generated the PD subject data collection forms. EGN analyzed the data. EGN wrote the manuscript. Editing by EGN, JC, AC, RM, and SR.

## References

1. Pringsheim, T., Jette, N., Frolkis, A. & Steeves, T. D. The prevalence of Parkinson’s disease: a systematic review and meta-analysis. Mov. Disord. 29, 1583–1590 (2014).

2. Marras, C. et al. Prevalence of Parkinson’s disease across North America. NPJ Parkinsons Dis. 4, 21 (2018).

3. Lee, H. J. et al. Tremor frequency characteristics in Parkinson’s disease under resting-state and stress-state conditions. J. Neurol. Sci. 362, 272–277 (2016).

4. Lee, S. Y. et al. Activities of daily living questionnaire from patients’ perspectives in Parkinson’s disease: a cross-sectional study. BMC Neurol. 16, 73 (2016).

5. Salat, D. & Tolosa, E. Levodopa in the treatment of Parkinson’s disease: current status and new developments. J. Parkinsons Dis. 3, 255–269 (2013).

6. Moran, A., Bergman, H., Israel, Z. & Bar-Gad, I. Subthalamic nucleus functional organization revealed by parkinsonian neuronal oscillations and synchrony. Brain 131, 3395–3409 (2008).

7. Eusebio, A. et al. Deep brain stimulation can suppress pathological synchronisation in Parkinsonian patients. J. Neurol. Neurosurg. Psychiatry 82, 569–573 (2010).

8. Cools, R., Barker, R. A., Sahakian, B. J. & Robbins, T. W. L-Dopa medication remediates cognitive inflexibility, but increases impulsivity in patients with Parkinson’s disease. Neuropsychologia 41, 1431–1441 (2003).

9. Cyron, D. Mental side effects of deep brain stimulation (DBS) for movement disorders: the futility of denial. Front. Integr. Neurosci. 10, 17 (2016).

10. Hickey, P. & Stacy, M. Deep brain stimulation: a paradigm shifting approach to treat Parkinson’s disease. Front. Neurosci. 10, 173 (2016).

11. Buhmann, C. et al. Adverse events in deep brain stimulation: a retrospective long-term analysis of neurological, psychiatric and other occurrences. PLoS One 12, e0178984 (2017).

12. Anguera, J. A. et al. Video game training enhances cognitive control in older adults. Nature 501, 97–101 (2013).

13. Kadakia, K., Patel, B. & Shah, A. Advancing digital health: FDA innovation during COVID-19. NPJ Digit. Med. 3, 161 (2020).

14. Mirelman, A. et al. Addition of a non-immersive virtual reality component to treadmill training to reduce fall risk in older adults (V-TIME): a randomised controlled trial. Lancet 388, 1170–1182 (2016).

15. Canning, C. G. et al. Virtual reality in research and rehabilitation of gait and balance in Parkinson disease. Nat. Rev. Neurol. 16, 409–425 (2020).

16. Wingeier, B. et al. Intra-operative STN DBS attenuates the prominent beta rhythm in the STN in Parkinson’s disease. Exp. Neurol. 197, 244–251 (2006).

17. Kühn, A. A. et al. High-frequency stimulation of the subthalamic nucleus suppresses oscillatory beta activity in patients with Parkinson’s disease in parallel with improvement in motor performance. J. Neurosci. 28, 6165–6173 (2008s).

18. Nabi, A., Mirzadeh, M., Gibou, F. & Moehlis, J. Minimum energy desynchronizing control for coupled neurons. J. Comput. Neurosci. 34, 259–271 (2012).

19. Wilson, D. & Moehlis, J. Clustered desynchronization from high-frequency deep brain stimulation. PLoS Comput. Biol. 11, e1004673 (2015).

20. Rosin, B. et al. Closed-loop deep brain stimulation is superior in ameliorating Parkinsonism. Neuron 72, 370–384 (2011).

21. Little, S. et al. Adaptive deep brain stimulation in advanced Parkinson disease. Ann. Neurol. 74, 449–457 (2013).

22. Adamchic, I. et al. Coordinated reset neuromodulation for Parkinson’s disease: proof-of-concept study. Mov. Disord. 29, 1679–1684 (2014).

23. Syrkin-Nikolau, J. et al. Coordinated reset vibrotactile stimulation shows prolonged improvement in Parkinson’s disease. Mov. Disord. 33, 179–180 (2017).

24. Usrey, W. M. & Alitto, H. J. Visual functions of the thalamus. Annu. Rev. Vis. Sci. 1, 351–371 (2015).

25. Sieben, K., Röder, B. & Hanganu-Opatz, I. L. Oscillatory entrainment of primary somatosensory cortex encodes visual control of tactile processing. J. Neurosci. 33, 5736–5749 (2013).

26. Bieler, M., Xu, X., Marquardt, A. & Hanganu-Opatz, I. L. Multisensory integration in rodent tactile but not visual thalamus. Sci. Rep. 8, 15684 (2018).

27. Koller, W. C., Vetere-Overfield, B. & Barter, R. Tremors in early Parkinson’s disease. Clin. Neuropharmacol. 12, 293–297 (1989).

28. Dirkx, M. F., Zach, H., Bloem, B. R., Hallett, M. & Helmich, R. C. The nature of postural tremor in Parkinson disease. Neurology 90, e1095–e1103 (2018).

29. Ackmann, J. J., Sances, A., Jr., Larson, S. J. & Baker, J. B. Quantitative evaluation of long-term Parkinson tremor. IEEE Trans. Biomed. Eng. 24, 49–56 (1977).

30. Percival, D. P. On estimation of the wavelet variance. Biometrika 82, 619–631 (1995).

31. Sanderson, J., Sudoyo, H., Karafet, T. M., Hammer, M. F. & Cox, M. P. Reconstructing past admixture processes from local genomic ancestry using wavelet transformation. Genetics 200, 469–481 (2015).

32. Malling, A. S. B. et al. The influence of posture duration on hand tremor during tasks with attention-distraction in persons with Parkinson’s disease. J. Neuroeng. Rehabil. 16, 61 (2019).

33. Jankovic, J., Schwartz, K. S. & Ondo, W. Re-emergent tremor of Parkinson’s disease. J. Neurol. Neurosurg. Psychiatry 67, 646–650 (1999).

34. Alty, J., Cosgrove, J., Thorpe, D. & Kempster, P. How to use pen and paper tasks to aid tremor diagnosis in the clinic. Pract. Neurol. 17, 456–463 (2017).

35. Allone, C. et al. Cognitive impairment in Parkinson’s disease, Alzheimer’s dementia, and vascular dementia: the role of the clock-drawing test. Psychogeriatrics 18, 123–131 (2018).

36. Koch, E. W. & Rosolowsky, E. W. Filament identification through mathematical morphology. Mon. Not. R Astron. Soc. 452, 3435–3450 (2015).

37. Clark, S., Peek, J. & Putman, M. Magnetically aligned HI fibers and the rolling Hough transform. Astrophy. J. 789, 82 (2014).

38. Gowers, W. R. A manual of diseases of the nervous system. by W. R. Gowers, M.D., F.R.C.P. J. and A. Churchill, 1886, Vol. I. J. Ment. Sci. 32, 594–596 (1887).

39. Ritto, A. P., Juste, F. S., Stuart, A., Kalinowski, J. & De Andrade, C. R. Randomized clinical trial: the use of SpeechEasy<sup>®</sup> in stuttering treatment. Int. J. Lang. Commun. Disord. 51, 769–774 (2016).

40. Gallop, R. F. & Runyan, C. M. Long-term effectiveness of the SpeechEasy fluency-enhancement device. J. Fluency Disord. 37, 334–343 (2012).

41. Raethjen, J. et al. Provocation of Parkinsonian tremor. Mov. Disord. 23, 1019–1023 (2008).

42. Roze, E. et al. Head tremor in Parkinson’s disease. Mov. Disord. 21, 1245–1248 (2006).

43. Sutton, G. G. & Sykes, K. The effect of withdrawal of visual presentation of errors upon the frequency spectrum of tremor in a manual task. J. Physiol. 190, 281–293 (1967).

44. Riley, P. O. & Rosen, M. J. Evaluating manual control devices for those with tremor disability. J. Rehabil. Res. Dev. 24, 99–110 (1987).

45. Riviere, C. & Thakor, N. Modeling and canceling tremor in human-machine interfaces. IEEE Eng. Med. Biol. Mag. 15, 29–36 (1996).

46. Gonzalez, J. G., Heredia, E. A., Rahman, T., Barner, K. E. & Arce, G. R. Optimal digital filtering for tremor suppression. IEEE Trans. Biomed. Eng. 47, 664–673 (2000).

47. Vescio, B. et al. Development and validation of a new wearable mobile device for the automated detection of resting tremor in Parkinson’s disease and essential tremor. Diagnostics (Basel) 11, 200 (2021).

48. Robertson, D. G. & Dowling, J. J. Design and responses of Butterworth and critically damped digital filters. J. Electromyogr. Kinesiol. 13, 569–573 (2003).

49. Meigal, A. Y. et al. Linear and nonlinear tremor acceleration characteristics in patients with Parkinson’s disease. Physiol. Meas. 33, 395–412 (2012).

