## Supplemental file for "Algorithmic Virtual Reality Reduces Parkinsonian Tremor"

### Supplemental Figure legends

**Supplemental Fig. 1. Parkinsonian tremor is observable with VR hardware.** Oculus Rift + Touch controller sensory hardware together with Unity software were utilized to record positional data associated with Parkinsonian IHT. Data was captured at 4Hz of changes in rotational angles $\alpha, \beta, \gamma$ (**a-c**) and Euclidian axes X, Y, Z (**d-f**). Data of the physical position of controllers is displayed in green and represent subject hands in Classical Reality (CR) (i.e., real-world). Data of the real-time position of study subjects' digital hands in VR is displayed in yellow. Parkinsonian tremors were more readily observable with rotational data. Data shown is from a tremor dominant hand of a single representative PD subject performing a PTT. Low amplitude changes in X, Y, Z coincident with rotational changes were observed in traces of some other subjects (data not shown) indicating recording in these dimensions is possible.

**Supplemental Fig. 2. Qualitative evidence of Classical Reality (i.e., real-world) parkinsonian tremor reduction associated with VR tremor stabilization.** PD subjects were observing their digital hands in VR with tremors (TSA-OFF) or without tremors (TSA-ON). Data here is of CR changes in subjects hand tremors (i.e., real physical world changes). Representative $\beta$ rotational changes of TSA-OFF are blue and TSA-ON are green. Qualitative assessment of these traces indicated that some subjects exhibited tremors in CR regardless of TSA ON or OFF. Representative subject 8 in (**a**). However, subject 5 had notably reduced tremors during TSA-ON (**b**).

**Supplemental Fig. 3. Simulation of tremor-related parkinsonian drawing irregularities and feature extraction with FilFinder.** Lines (**a**) were seeded with increasing amounts of salt and pepper noise (**d**, **g**, and **j**) to simulate the effect of hand articulation of brush strokes with parkinsonian tremor. FilFinder detected line features (red overlay in **b**, **e**, **h**, and **k**). And converted them to one-pixel wide filaments (**c**, **f**, **i**, and **l**). Salt and pepper noise was generated by drawing random numbers using the number generator in python's numeric library. Arbitrary values inputted to the number generator to progressively distort lines and create micro-curvatures were 0, 0.0125, 0.025, and 0.05 for **a**, **d**, **g** and **j**, respectively.

**Supplemental Fig. 4. Quantification of simulated parkinsonian tremor irregularities.** Rolling Hough Transform was applied to one-pixel wide filaments extracted by FilFinder (see Supplemental Fig. 3) to obtain the distribution of angular values of microcurvatures. These are portrayed in $2_\theta$ radar charts (**a-d**). The percent area of these $2_\theta$ charts is plotted against the amount of simulated tremor (**e**). Micro-curvatures were dose-dependently related to the amount of simulated tremor. Percent area was defined as the gray region in $2_\theta$ charts relative to the entire $2_\theta$ chart. Values for simulated tremors are same as in Supplemental Fig. 3. n = 5 for each simulated tremor value.

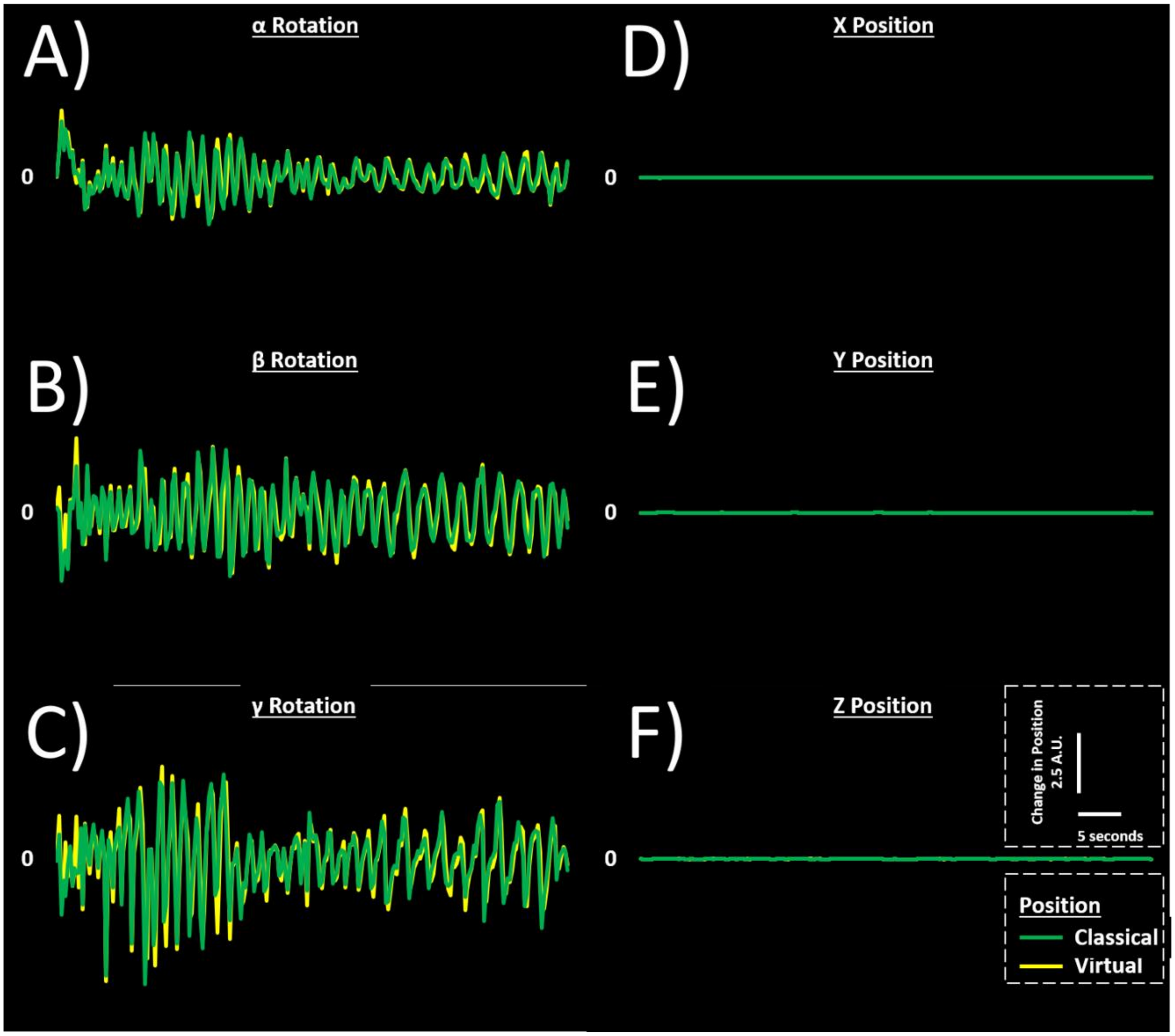


**Supplemental Figure 1**

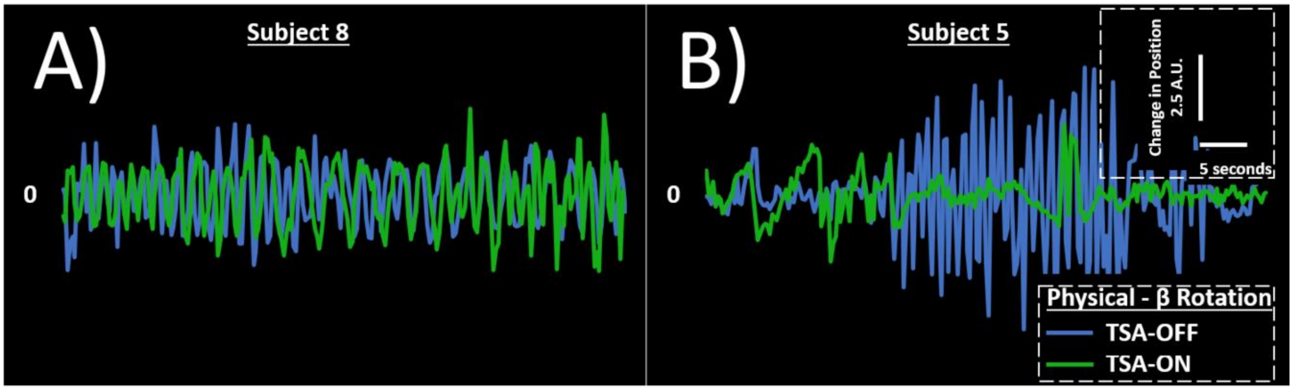


**Supplemental Figure 2**

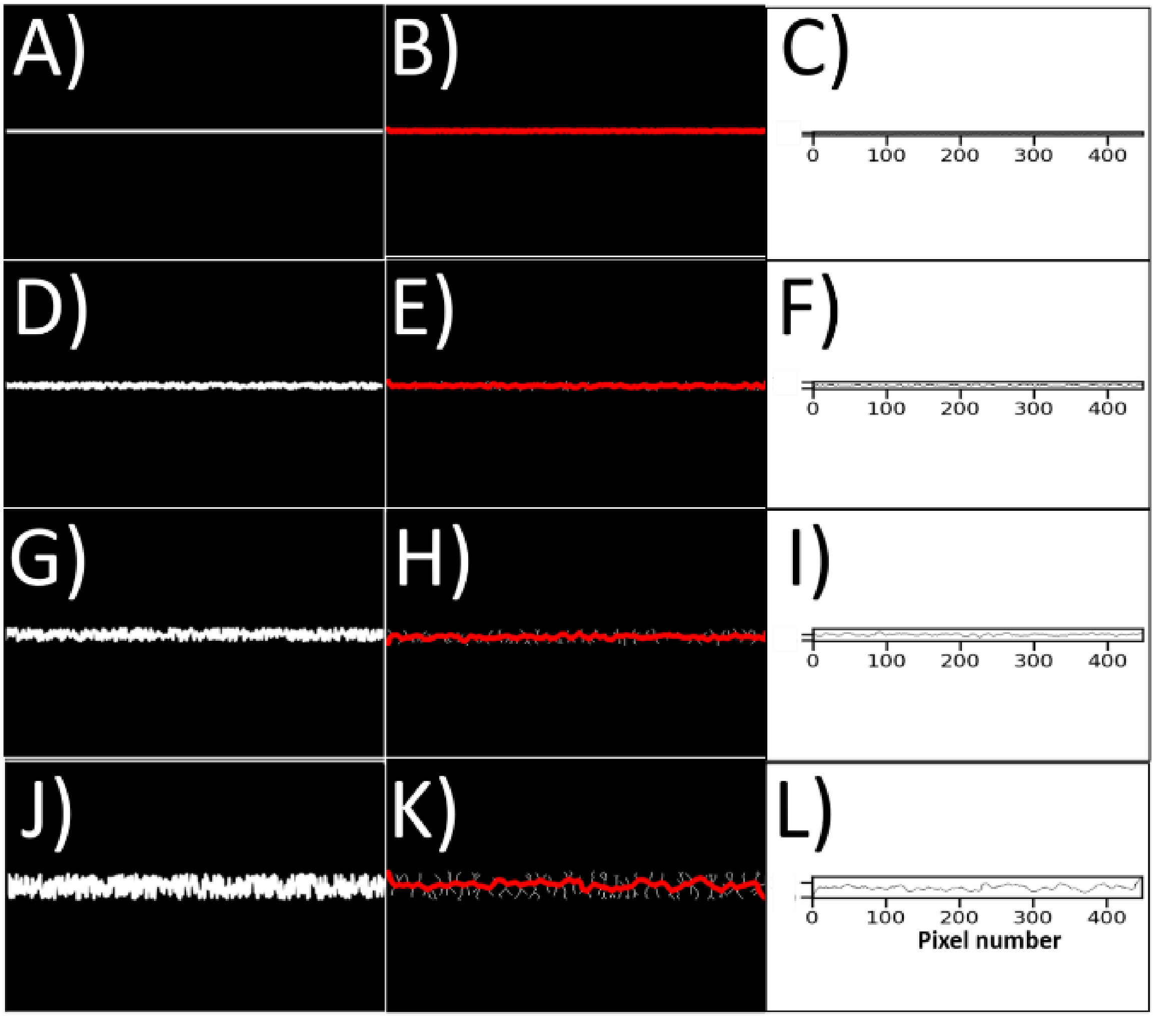


**Supplemental Figure 3**

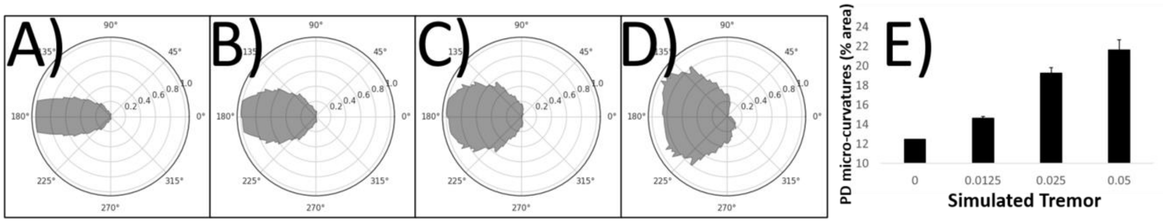


**Supplemental Figure 4**
